# Waning of post-vaccination neutralizing antibody responses against SARS-CoV-2, a systematic literature review and meta-analysis

**DOI:** 10.1101/2023.08.08.23293864

**Authors:** Henning Jacobsen, Ioannis Sitaras, Maeva Katzmarzyk, Viviana Cobos Jiménez, Robert Naughton, Melissa M. Higdon, Maria Deloria Knoll

## Abstract

**Background:** Mass COVID-19 vaccination and the continuous introduction of new viral variants of SARS-CoV-2, especially of Omicron subvariants, has resulted in an increase in the proportion of the population with hybrid immunity at various stages of waning protection. We systematically reviewed waning of post-vaccination neutralizing antibody titers in different immunological settings to investigate potential differences.

**Methods:** We searched for studies providing data for post-vaccination neutralizing antibody responses against SARS-CoV-2 in PubMed, bioRxiv, and medRxiv from Dec 15, 2021, to Jan 31, 2023, using keywords related to COVID-19, vaccination, and antibody neutralization. We used random effects meta-regression to estimate the average fold-reduction in post-vaccination neutralizing antibody titers against the Index strain or Omicron BA.1. from month 1 to month 6 post last dose, stratified by vaccination regimen (primary or booster) and infection-naïve vs hybrid-immune status.

**Findings:** In total, 26 studies reporting longitudinal post-vaccination neutralizing antibody titers were included. Neutralization titers against the Index variant were available from all studies for infection-naïve participants, and from nine for hybrid-immune participants. Against Omicron BA.1, nine and eight studies were available for infection-naïve and hybrid-immune cohorts, respectively. In infection-naïve cohorts, post-vaccination neutralization titers against the Index strain waned 5.1-fold (95% CI 3.4-7.8) from month 1 to month 6 following primary regimen and 3.8-fold (95% CI 2.4-5.9) following the booster. Titers against Omicron BA.1 waned 5.9-fold (95% CI 3.8-9.0) in infection-naïve, post-booster cohorts. In hybrid-immune, post-primary vaccination cohorts, titers waned 3.7-fold (95% CI 1.7-7.9) against the Index strain and 5.0-fold (95% CI 1.1-21.8) against Omicron BA.1.

**Interpretation:** No obvious differences in waning between post-primary or post-boost vaccination were observed for vaccines used widely to date, nor between infection-naïve and hybrid-immune participants. Titers against Omicron BA.1 may wane faster compared to Index titers, which may worsen for more recent Omicron sub-variants and should be monitored. Relatively small datasets limit the precision of our current analysis; further investigation is needed when more data become available. However, based on our current findings, striking differences in waning for the analyzed and future comparisons are unlikely.

## Introduction

COVID-19 vaccines continue to effectively protect against severe disease and death caused by SARS-CoV-2 despite continuous viral evolution and waning immunity^1–3^. However, vaccine effectiveness against SARS-CoV-2 transmission, infection, and symptomatic disease has declined, and immunity against the Wuhan Index strain, either elicited by vaccination or previous infection, shows little protection against infection with Omicron-related viral variants^2–4^. Thus hybrid immunity (immunity developed through a combination of SARS-CoV-2 infection and vaccination) involving infections with more recent viral variants is increasingly relevant. Clinical studies are imperative for assessing the impact of novel viral variants on vaccine performance and understanding the waning of protection after vaccination and/or infection, but these studies demand significant time. While laboratory data, such as neutralizing antibody titers, can be generated and shared much more quickly thereby potentially informing vaccine policy when clinical data are lacking, single studies often lack the power to provide sound and robust conclusions regarding complex biological functions such as antibody waning^5^. Meta-analysis of data across studies can increase power and can evaluate impact of different immunological factors, including number of doses and effects of hybrid immunity.

We systematically reviewed the evidence of post-vaccination neutralization antibody titers against the Index strain and Omicron BA.1 over time and compared the degree of waning after the last dose between primary and booster vaccination and between infection-naïve and hybrid-immune participants.

## Methods

### Search strategy and selection criteria

The systematic review and meta-regression were conducted according to the Preferred Reporting Items for Systematic Reviews and Meta-Analyses (PRISMA) guidelines.

We searched PubMed, medRxiv, and bioRxiv from December 15, 2021, to January 31, 2023, using the keywords “COVID-19”, “Omicron”, and “neutralization”. Two reviewers (HJ, IS) screened titles and abstracts and conducted full-text review; inclusion was limited to studies providing neutralization data against both the Index (Wuhan-line) strain and Omicron BA.1.

To investigate systematically if booster doses (compared to primary series) or hybrid immunity (compared to infection naïve) affect the rate of neutralizing antibody waning, we performed meta-regressions assessing change in neutralization titers over time for strata with six or more cohorts. We included studies reporting post-vaccination neutralizing antibody titers (using authentic virus or pseudo-virus neutralization assays) for at least two time-points following last vaccine dose. In the case of pseudo-virus neutralization assays, we included only studies where the pseudo-viruses used carried the complete complement of spike mutations characteristic of the variant they represented. Data resulting from surrogate neutralization titers were not assessed. Per study, all cohorts were assessed that matched the inclusion criteria and, therefore, one study could contribute multiple observations from different cohorts. We collected outcomes from studies investigating infection-naïve cohorts post-primary or post first booster vaccination, and from hybrid-immune cohorts, post-primary vaccination. Studies were excluded if they did not provide neutralization titers against the Index strain and Omicron BA.1, did not provide neutralization data for at least two time points, if cohort characteristics did not match the scope of the analysis (assessment of non-licensed vaccines, immunocompromised participants) or if information regarding previous infection history of the study cohort was insufficient. We excluded studies evaluating infection-naïve, post-primary vaccination titers against Omicron BA.1 because of overall low or undetectable titers^6,7^. Geometric mean titers (GMT) against the Index strain and Omicron BA.1 as measured by authentic virus neutralization assays or pseudo-virus-based neutralization assays were abstracted.

### Assessment of study reliability

We systematically assessed the reliability of included studies using a tool we previously developed tailored for studies reporting post-vaccination neutralizing antibody responses^5^. The tool assesses reporting quality (e.g., methodological detail, description of relevant clinical data, etc.), overall strength of the data, and standardization measures using a standardized set of criteria and metrics. Each aspect is rated with an output (no, low, medium, high, or unclear risk of unreliability), resulting in an overall score for each study.

### Analysis

Average declines in GMTs were estimated stratified by dose (primary, first booster), prior infection status (naïve vs hybrid-immune), and strain (Index vs. Omicron BA.1). Primary vaccination was defined as one dose of Ad26.CoV2.S vaccine or two doses of any other included vaccine. Booster vaccination was defined as one dose of any COVID-19 vaccine after any primary series vaccination. Hybrid-immune cohorts included convalescent participants who had an infection prior to the last dose.

The natural log of neutralization antibody response GMT (logGMT) was calculated for all available time points post final dose. If not provided, GMT was calculated using raw data when available or abstracted from high-resolution figures. If the standard deviation (SD) corresponding to each logGMT was not provided, it was derived from confidence intervals (CIs); if no CIs were provided, within each of the five comparison groups, SD was imputed by calculating the median SD among other observations with SDs reported.

The average change in logGMT was estimated using a linear mixed effects model for the repeated measures within each comparison group (PROC MIXED; SAS 9.4). The standard errors calculated from SDs, and sample sizes abstracted from the studies were squared to produce estimates of residual variances for inverse weighting in the linear mixed effects model. The logGMT was regressed on months since vaccination; we evaluated non-linearity by including a quadratic term for time, which was not statistically significant in any model (all p > .15). Models were adjusted for vaccine platform. Difference in degree of waning by dose, prior infection status, strain, or vaccine platform was assessed using an interaction term with time in models, adjusting for other covariates. Confidence interval bands for average logGMT over time in plots were estimated by re-defining the intercept in the model by centering the time variable monthly from 1 to 6 months. Results are presented as GMTs by exponentiating model outputs. Statistical significance was defined as p < .05; adjustments for multiple comparisons were not made.

## Results

We screened titles and abstracts from 8418 articles, of which 347 underwent full-text review and 26 were eligible for analyses (Figure 1). Abstracted neutralization titers and relevant cohort characteristics including study population, number of doses, vaccine product, and infecting strain are provided in Supplementary appendix 1.

**Figure 1:**
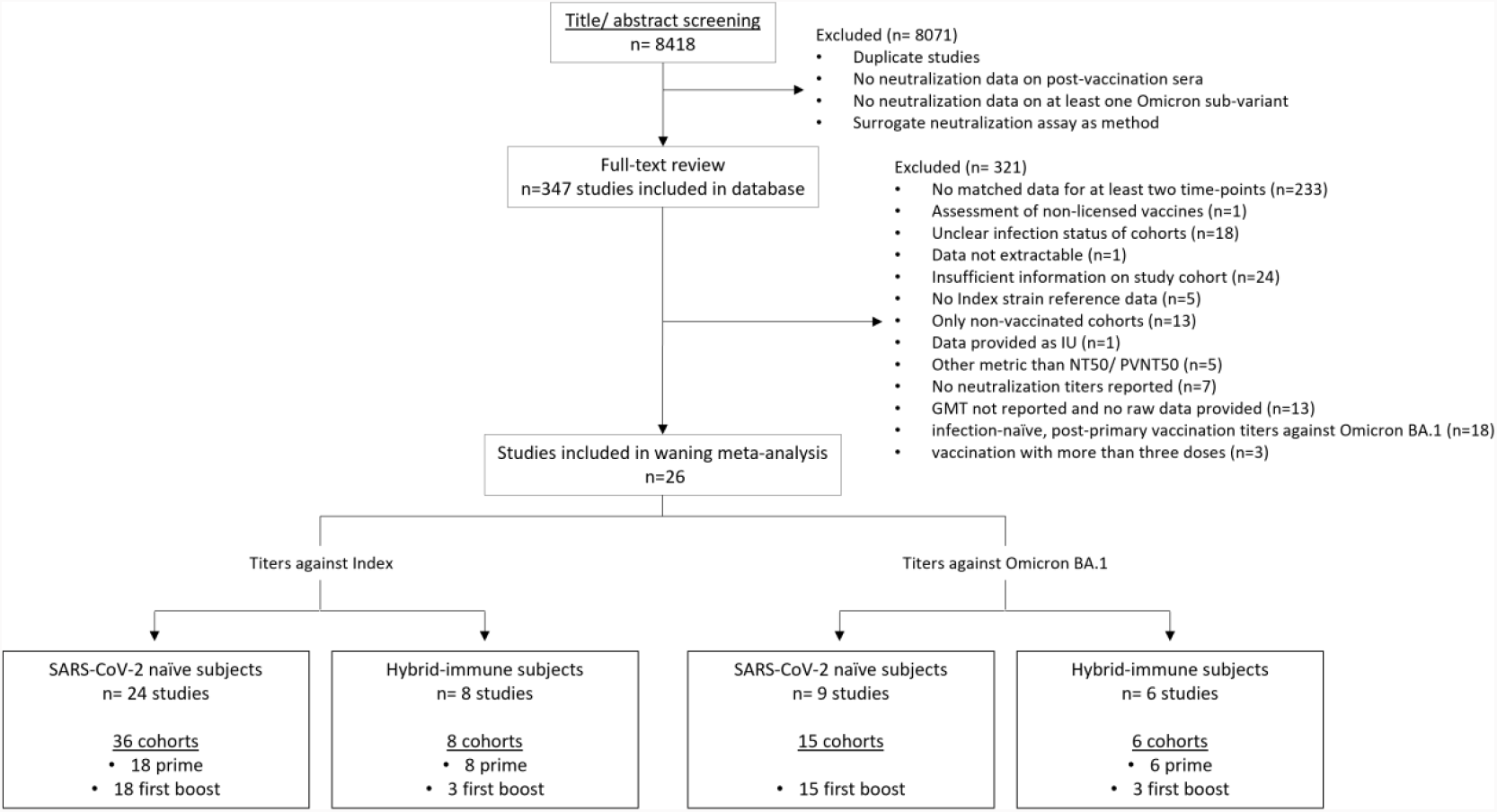
Study identification and Selection.

Five strata had six or more cohorts for meta-analyses: 1) infection-naïve, post-primary vaccination titers against the Index strain (n= 18 cohorts); 2) infection-naïve, post-boost vaccination titers against the Index strain (n= 18); 3) infection-naïve, post-boost vaccination titers against Omicron BA.1 (n= 15); 4) hybrid-immune, post-primary vaccination against the Index strain (n= 8); and 5) hybrid-immune, post-primary vaccination against Omicron BA.1 (n= 6; Figure 1). Too few (≤ 3 cohorts) assessed hybrid-immune, post-booster vaccination titers, vaccination with four or more doses, or vaccination with variant-adapted vaccines, and were therefore not meta-analyzed. Among hybrid-immune cohorts, all studies evaluated infections occurring prior to the last dose except one, which provided data after breakthrough infection but was excluded from meta-analysis because sampling time-points were unclear^8^]. All hybrid-immunity cohorts were from pre-Omicron infections so we could not assess impact of variant-specific effects on hybrid-immunity.

There was wide heterogeneity in peak GMTs across studies within strata, for example ranging between 101 and 4096 among infection-naïve participants boosted with mRNA vaccines (Figure 2b, supplementary appendix 1), resulting in wide confidence intervals of meta-analyses. Average peak GMTs differed between strata, with highest average GMTs observed against the Index strain in hybrid-immune post-primary participants and lowest against Omicron BA.1 in infection-naïve post-boost participants (Table 1, Figure 2). As expected, average peak titers post-vaccination were higher in subjects with an infection history compared to naïve subjects and titers against Omicron BA.1 were generally lower than those against the Index strain. Mean peak titers were significantly higher for mRNA vaccines compared to other vaccine platforms; however, few studies evaluated inactivated (n=3 cohorts from 3 studies^9–11^) or viral vector vaccines (n= 8 cohorts from 6 studies^12–17^), and only two of these studies provide direct comparisons to other platforms^12,13^.

**Table 1:** Average peak and 5-month waning of neutralizing antibody titers in infection-naïve and hybrid-immune cohorts against the Index strain and Omicron BA.1.

| Infection status | Dose | Strain | Group | Group size (studies/cohorts/Participants) | Average Peak GMT (95% CI) | mRNA cohorts [%] | Average fold decline in GMT, 1-6 months (95% CI) |
| --- | --- | --- | --- | --- | --- | --- | --- |
| Infection naïve | Prime | Index | A | 16/19/438 | 718.0 (384.0-1341.0) | 61.1 | 5.1 (3.4-7.8) |
|  | Boost | Index | B | 13/21/620 | 954.0 (466.0-1951.0) | 50.0 | 3.8 (2.4-5.9) |
|  |  | Omicron BA.1 | D | 10/16/584 | 169.0 (76.0-377.0) | 53.3 | 5.9 (3.8-9.0) |
| Hybrid-immune | Prime | Index | C | 7/8 /143 | 2803.0 (1651.0-4760.0) | 62.5 | 3.7 (1.7-7.9) |
|  |  | Omicron BA.1 | E | 6/6/93 | 339.0 (193.0-595.0) | 50.0 | 5.0 (1.1-21.8) |

**Figure 2:**
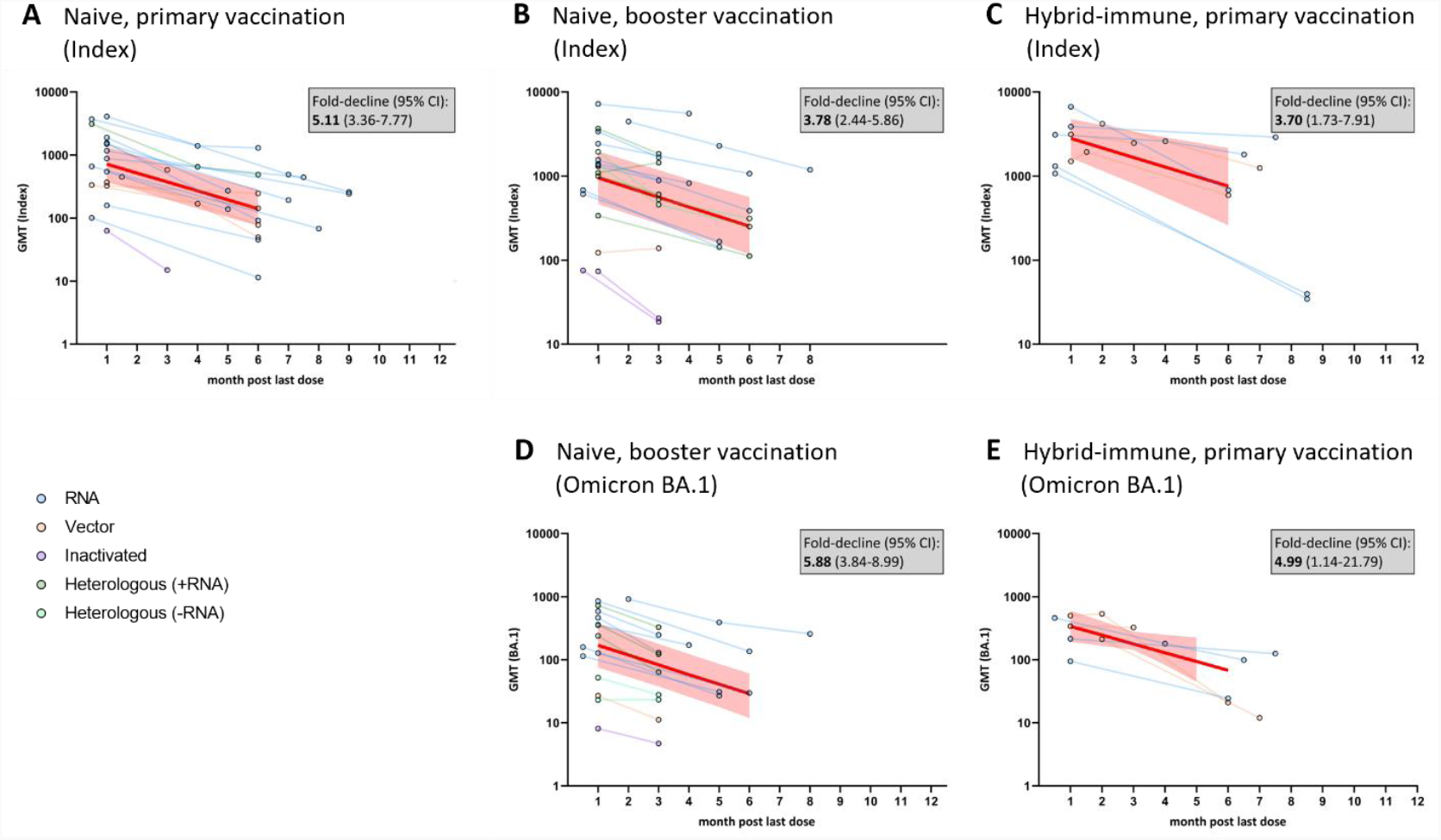
Neutralizing antibody titers over time since last vaccination against the Index strain or Omicron BA.1 in infection-naïve or hybrid-immune participants after primary or booster vaccination. Rates of waning against the Index strain (A – C) and against Omicron BA.1 (D and E) are shown stratified by prior infection status and dose. Lines connecting data points represent individual cohorts, color coded by vaccine platform. Bold red lines represent average declines from meta-regression for each stratum; shaded area represents 95% confidence intervals of GMT over time. Abbreviations: GMT, geometric mean titer; CI, confidence interval; Index, SARS-CoV-2 Wuhan-like including D614G-strains; +mRNA, heterologous vaccine regimen involving at least one mRNA-vaccine dose; -mRNA, heterologous vaccine regimen involving no mRNA-vaccine dose.

Average neutralization titers declined from month 1 to 6 in all five strata, ranging from 3.7-fold (95%CI 1.7-7.9) against the Index strain in hybrid-immune participants post-primary to 5.9 (95% CI 3.8-9.0) against Omicron BA.1 in infection-naïve boosted participants (Table 1, Figure 2), but the declines were not statistically significantly different between strata (p= .67). The rate of waning in the first 6 months appeared linear in all five strata (all p > 0.15 for quadratic term), but most cohorts (39 of 48; 81.2%) provided data for only two time-points and no eligible studies had more than three time-points. Although declines in neutralizing antibodies cross-reactive to Omicron BA.1 appeared to be greater than declines of Index-specific responses in both infection-naïve and hybrid-immune cohorts (5.0-to 5.9-fold reductions vs. 3.7-to 3.8-fold, respectively), they were not statistically significant (p= .22), nor were rates of decline statistically different for any covariate evaluated (all p > .17).

Neutralization titers declined in all cohorts except two (Figure 2 panel B), that were from a single study^12^ that assessed titers at shorter follow-up times (months 1 and 3), among participants vaccinated with a vector vaccine (of five comparable cohorts that evaluated vector vaccines). Statistically significant increases in neutralizing antibody (GMTs from 1090.5 to 1444.3) against the Index strain were observed in one cohort that received a heterologous booster with Ad26.CoV2.S as a third dose after two doses of mRNA vaccine in infection-naïve participants. There was no change in a second cohort that received two doses of Ad26.CoV2.S (GMTs 122.8 and 138.2); however, titers against Omicron BA.1 declined in both (358.9 to 123.0 and 26.9 to 11.2, respectively; both p < .05) and significant declines were observed for two other cohorts in the same study that received either mRNA vaccines or heterologous vaccination with Ad26.CoV2.S as a first dose.

### Assessment of study reliability

Assessment of reliability of the 26 eligible studies classified only four studies (15.4%) as having high reliability; four (15.4%) had medium reliability, six (23.1%) had low reliability, and 14 (53.9%) had unclear reliability because critical information was not provided (Supplementary Figure 1). Unclear or low-reliability scores were primarily attributable to poor reporting quality (e.g., input titer used, spike complement, etc.) regarding pseudo-virus constructs (seven studies, 26.9%) or assay standardization (12 studies, 46.2%; Supplementary Figure 1). Analyses stratified by reliability score showed that neither peak titers nor waning rates differed markedly between studies with medium to high reliability scores compared to low reliability (Supplementary Figure 2). Individual scoring results are provided as Supplementary Appendix 2.

## Discussion

Through a systematic literature review and meta-analysis, we found neutralizing antibodies declined after COVID-19 vaccination from months one to six ranging from 3.7-fold to 5.9-fold when evaluating post-primary or first booster against either the Index strain or Omicron BA.1. Waning rates were generally similar after primary or first booster regimens, and between infection-naïve and previously infected cohorts. Declines of neutralizing antibodies cross-reactive to Omicron BA.1 were greater than declines of Index-specific responses, both in infection-naïve and hybrid-immune cohorts, though this difference was not statistically significant. Only three studies evaluated a second booster; two reported no significant differences in waning kinetics between first and second booster^26,27^ and one reported slightly enhanced antibody durability after the second booster, but the second booster cohort was small (n=7)^21^. Because waning was similar after primary and first booster doses, degree of waning with subsequent doses is also expected to be similar. However, data to assess long-term waning, such as 12 months after the last vaccine dose, were unavailable, complicated both by needing to wait that long and by study subjects getting revaccinated before that time. As duration between vaccinations increases, this may be addressed in future studies. These waning rates could be used to predict waning for future relevant scenarios and adapt vaccination strategies accordingly.

Declines in neutralization titers were observed in all but three infection-naïve cohorts evaluated. One cohort that received a heterologous boost regimen (inactivated prime followed by a vector boost) was followed for 3 months and no change in titers was observed^9^. However, overall titers were low throughout. Two additional cohorts, one vaccinated with two doses of vector vaccine (out of five available vector-immunized cohorts) and one with a heterologous regimen (mRNA prime followed by a vector boost) had titers that increased against the Index strain through month 3 (longer follow-up was not conducted)^12^. Titers against Omicron BA.1 declined in these cohorts indicating that no undetected breakthrough infection occurred driving these titer increases. Interestingly, these three exceptional cohorts all received a vector vaccine as the last dose and hence it can be speculated that vector-mediated immunization might cause more durable antibody responses early after immunization/booster. On the other hand, studies with longer-term follow-up support overall comparable rates of waning across vaccine platforms beyond three months after the last dose, which might be explained by full clearance of the vector and any benefit it might add. More studies are needed to address this important observation and explore the potential role vector vaccines could play in enhancing durable immune responses. While we and others have shown that vector vaccines are generally less immunogenic compared to mRNA vaccines, heterologous regimens combining mRNA and vector vaccines have been shown to elicit immune responses comparable to mRNA vaccinations alone^13,29^. Hence, boosters with vector vaccines in mRNA-primed (or already mRNA-boosted) participants could elicit the highest and most durable immune responses. This has been shown by Lyke et al. who observed that titers were more stable in subjects who received an mRNA prime followed by a vector boost than subjects who received three doses of mRNA vaccine^12^. We identified four additional cohorts (from three studies) evaluating hybrid-immune subjects that did not show a significant decline of titers in the observed period, and two of these cohorts were followed for more than six months post-last vaccine dose^8,16,30^. However, two of the studies did not investigate breakthrough infections after the last vaccine dose^8,30^. Breakthrough infections in even a small proportion of the subjects can have a large impact on the overall GMT because the impact of these few infections on the overall GMT of a group can be large. Therefore, these studies should be considered with caution. Still, one study on hybrid-immune subjects that ruled out breakthrough infections after the last vaccine dose showed stable titers over a three-month period^16^ which supports other observations that hybrid-immunity might have the potential to stabilize antibody titers at least temporarily^21^.

Neutralizing antibody titers can support and complement clinical vaccine effectiveness data as they correlate well with protection against infection and mild disease. Even if neutralizing antibodies fail to hinder initial infection and symptomatic disease, they will limit initial viral load and thus mitigate disease progression, so they correlate also with protection against severe disease. This can be seen by the see-saw pattern of COVID-19 vaccine effectiveness against severe disease which was similar to titers peaking in the first weeks after each dose and falling thereafter until the next dose^3^. However, the observed larger declines in titers against Omicron relative to the Index strain may correlate less well to clinical vaccine effectiveness against severe disease, which shows less waning than VE against symptomatic disease and infection. This supports that protection is aided by additional factors such as cellular immunity, which has gained increasing recognition for its importance for protection against severe disease^18^. While no precise correlates of protection are defined for neutralizing antibodies, an understanding of overall titers and waning rates will allow us to predict how fast protection against infection and mild disease will decline and whether this might differ by vaccine type, regimen, infection history or characteristics such as sex, age, or comorbidities to inform vaccine policy, including the time interval when additional vaccine doses should be given.

But even studying neutralization titers takes time; very few longitudinal studies with data against Omicron subvariants other than for BA.1 were available at the time of this review, and increasing immune escape from post-vaccination neutralizing antibody responses resulting in large proportions of participants with undetectable titers makes them difficult to evaluate, especially in infection-naïve cohorts post-primary vaccination. Indeed, available studies for newer sub-variants provide contradictory results with some observing increased rates of waning against Omicron sub-variants compared to the Index variant^19,20^, some finding similar rates of waning^21– 23^, and others reporting lower rates^24,25^. More evidence is needed to determine if waning of post-vaccination neutralizing antibody titers cross-reactive to newer Omicron subvariants differs from those reactive to the Index variant or to Omicron BA.1. Such results may depend on whether the vaccine targets the emerging subvariants. We found only one study assessing waning of neutralizing antibodies after bivalent mRNA vaccination (Index plus BA.4/5 antigen), which found greater waning during the first three months against Omicron subvariants than against the Index variant^28^, similar to our observations for monovalent Index-directed vaccines. We were also unable to assess variant-specific effects on hybrid-immunity since all hybrid-immune cohorts investigated involved pre-Omicron infections.

Our results confirm observations of superior immunogenicity of some vaccination strategies over others. We observed significant differences in overall titers by vaccine platform, with mRNA vaccines resulting in higher titers and inactivated vaccines the lowest. Importantly, waning rates were not significantly different between the platforms. These results support previous findings that both booster doses and hybrid-immunity significantly increase overall titers and titers against Omicron BA.1 are generally lower than against the Index strain^8,30–33^. Importantly, these results provide evidence of a relatively constant rate of waning for the different groups included in the analysis; thus, individuals immunized with a less immunogenic primary regimen are likely to reach non-protective antibody titers faster. This effect becomes more significant when comparing primary regimen to hybrid-immune or boosted cohorts. These results may prove informative for booster strategies, especially when vaccine supply is low or if over-immunization should be avoided because of possible imprinting and a lack of variant-adapted vaccines.

A systematic assessment of study reliability revealed that 88% of included studies had medium, low, or unclear reliability scores reflecting primarily poor reporting quality of study methods and details. While this does not necessarily translate to biased or unreliable data, the overall low-reliability scores and small percentage of studies with a high-reliability score reflects that data on neutralizing antibodies are difficult to compare across studies^4,5^. This finding is further reflected by the wide confidence intervals observed in our meta-regression results. However, we included all studies meeting inclusion criteria irrespective of their reliability score for sample size reasons. A sensitivity analysis did not find an association between study results and reliability score, i.e., poorer scores were not more likely to be outliers.

In summary, neutralizing antibody titers are an important correlate of protection against infection and continued monitoring of new vaccines as they become available and new variants as they emerge will provide important alert signals and information to help guide vaccination regimens in the future. While absolute values of neutralization titers continue to vary widely between studies, this evaluation across many cohorts provides confidence that large differences in waning rates after booster doses are unlikely between vaccines used widely to date. However, significantly different baseline titers by vaccine platform and number of doses and infections means titers will drop to non-functional levels sooner for some conditions, which is particularly important for public health policymaking. Additionally, we could not evaluate the most recent conditions, and waning against Omicron sub-variants other than BA.1 might be slightly faster. This should be monitored carefully, as well as waning against clinically relevant Omicron sub-variants after variant-adapted vaccination and the impact of hybrid-immunity by Omicron sub-variants, especially in combination with variant-adapted vaccination.

## Data Availability

All data produced in the present study are available upon reasonable request to the authors

## Contributors

HJ and IS performed the systematic literature review, screened, and excluded studies. HJ, MK, RN and IS abstracted data from included studies. HJ and IS calculated GMT, SD, SE, and 95% CI from raw data where necessary. MDK performed the meta-analysis. HJ performed initial interpretation and wrote the manuscript. HJ and MK prepared figures. IS, VCJ, DRF, MDK, RN and MH reviewed the manuscript and supported interpretation. MH acquired funding. All authors read and revised the manuscript.

## Acknowledgment

We thank William Dowling and Daniel Feikin for excellent discussion and for their input on interpreting the data. We thank the Peter und Traudl Engelhorn Foundation for providing a post-doctoral scholarship to H.J.

## Supplements

Supplementary appendix 1: Study characteristics and data from included studies

Supplementary appendix 2: Reliability assessment, individual assessment outcomes

Supplementary figure 1: Reliability assessment, summary

**Supplementary Figure 1:**
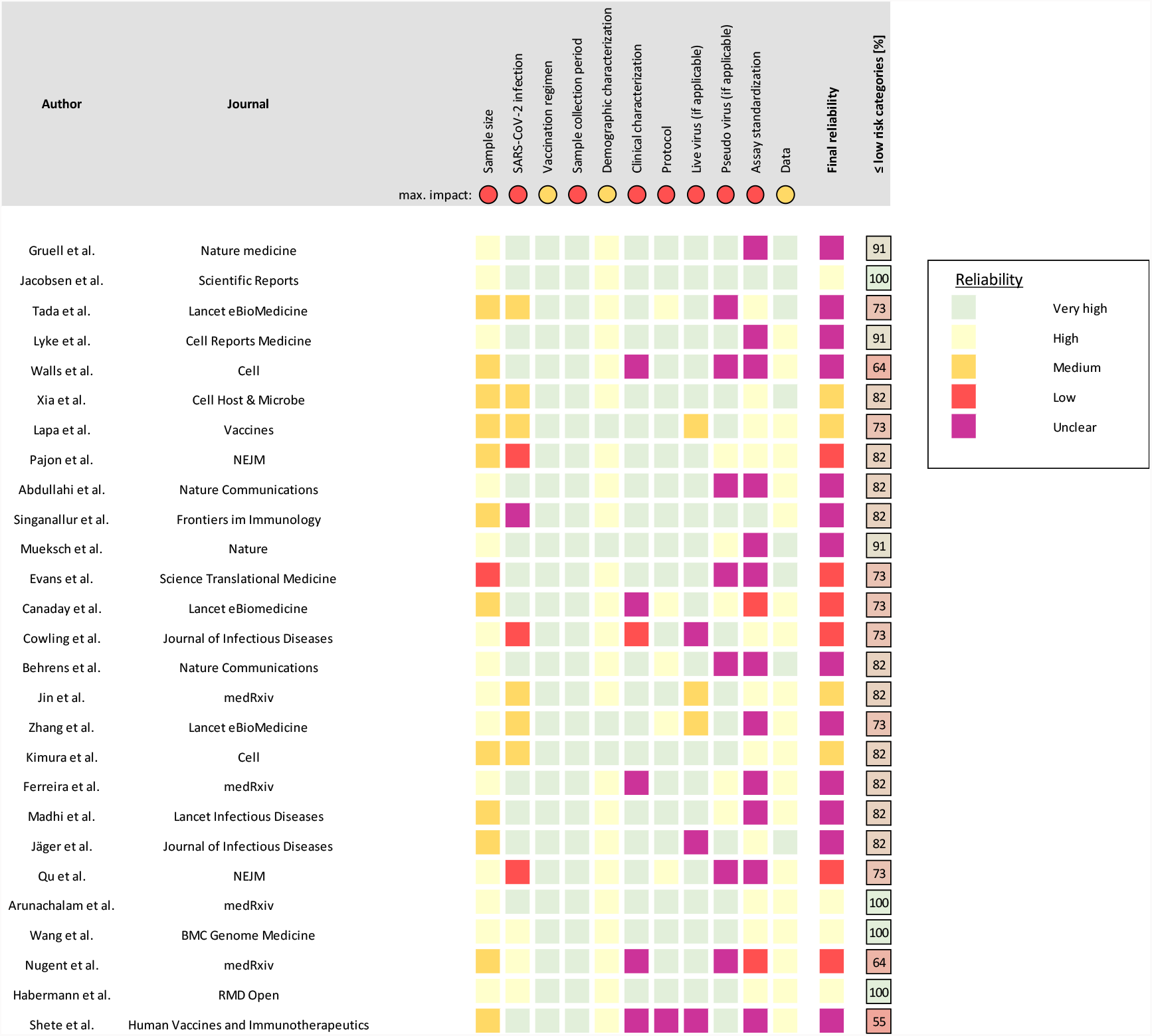
Reliability assessment. All studies that were included in the meta-analyses were assessed with a reliability score using a previously published tool^*5*^. Studies were considered to have high reliability if no criterion had more than a low risk score (yellow), medium reliability if no criterion was above a medium risk score (orange) and low if at least one criterion met a high risk score (red). No study met the “very high” reliability score (no criterion with risk of bias). Studies with at least one criterion that could not be assessed (e.g. no data provided or unclear), received an unclear reliability (purple). Eleven categories are assessed by the tool and assigned an independent risk score. The maximum impact a category can have is shown as “max impact” indicating the worst possible outcome for this category. The percentage of categories with a low or no risk is shown on the right, complementing the final reliability.

Supplementary figure 2: Reliability assessment, sensitivity analysis

**Supplementary Figure 2:**
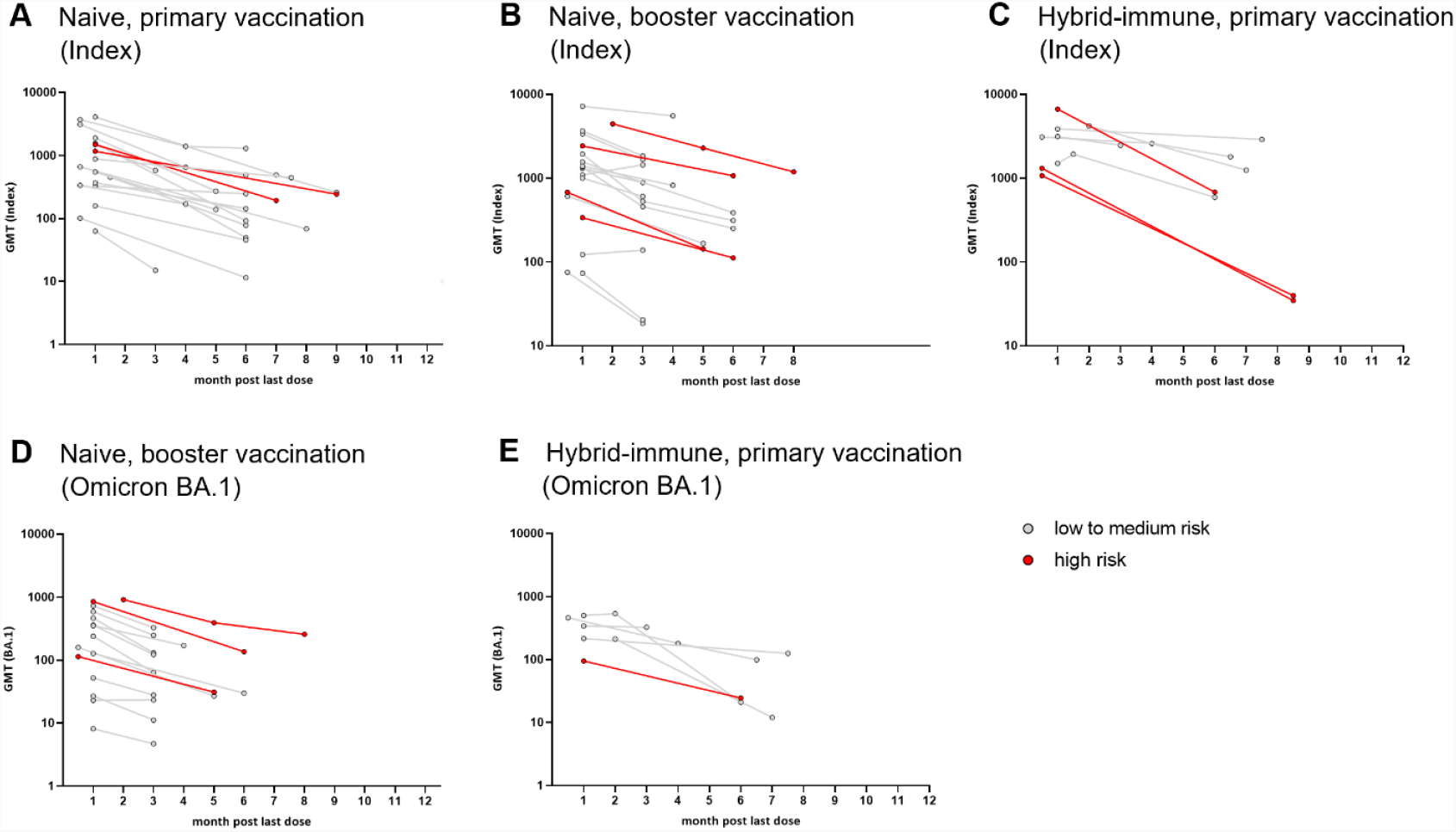
Reliability assessment of included studies. Rates of waning against the Index strain (A – C) and against Omicron BA.1 (D and E) shown colored according to reliability as assessed by a previously published standardized reliability assessment tool^*5*^. Studies assessed as high to medium reliability are shown in grey, low reliability studies are shown in red. Studies with unclear reliability are included to “low to medium risk”. Abbreviations: GMT, geometric mean titer; Index denotes SARS-CoV-2 Wuhan-like including D614G-strains.

## References

1. Higdon, M. M. et al. Duration of effectiveness of vaccination against COVID-19 caused by the omicron variant. Lancet Infect Dis 22, 1114–1116 (2022).

2. Bobrovitz, N. et al. Protective effectiveness of previous SARS-CoV-2 infection and hybrid immunity against the omicron variant and severe disease: a systematic review and meta-regression. Lancet Infect Dis S1473-3099(22)00801–5 (2023) doi:10.1016/S1473-3099(22)00801-5.

3. Feikin, D. R. et al. Assessing COVID-19 vaccine effectiveness against Omicron subvariants: Report from a meeting of the World Health Organization. Vaccine 41, 2329–2338 (2023).

4. Jacobsen, H. et al. Post-Vaccination Neutralization Responses to Omicron Sub-Variants. Vaccines (Basel) 10, 1757 (2022).

5. Jacobsen, H. et al. Assessing the Reliability of SARS-CoV-2 Neutralization Studies That Use Post-Vaccination Sera. Vaccines (Basel) 10, 850 (2022).

6. Wilhelm, A. et al. Limited neutralisation of the SARS-CoV-2 Omicron subvariants BA.1 and BA.2 by convalescent and vaccine serum and monoclonal antibodies. eBioMedicine 82, (2022).

7. Sitaras, I. et al. Systematic review of primary and booster COVID-19 sera neutralizing ability against SARS-CoV-2 omicron variant. NPJ Vaccines 7, 147 (2022).

8. Walls, A. C. et al. SARS-CoV-2 breakthrough infections elicit potent, broad, and durable neutralizing antibody responses. Cell 185, 872–880.e3 (2022).

9. Jin, L. et al. Antibody Persistence and Safety through 6 Months after Heterologous Orally Aerosolised Ad5-nCoV in individuals primed with two-dose CoronaVac previously. 2022.07.26.22278072 Preprint at https://doi.org/10.1101/2022.07.26.22278072 (2022).

10. Zhang, Y. et al. Immunogenicity, durability, and safety of an mRNA and three platform-based COVID-19 vaccines as a third dose following two doses of CoronaVac in China: A randomised, double-blinded, placebo-controlled, phase 2 trial. eClinicalMedicine 54, (2022).

11. Wang, X.-J. et al. Neutralization sensitivity, fusogenicity, and infectivity of Omicron subvariants. Genome Medicine 14, 146 (2022).

12. Lyke, K. E. et al. Rapid decline in vaccine-boosted neutralizing antibodies against SARS-CoV-2 Omicron variant. Cell Reports Medicine 3, 100679 (2022).

13. Behrens, G. M. N. et al. BNT162b2-boosted immune responses six months after heterologous or homologous ChAdOx1nCoV-19/BNT162b2 vaccination against COVID-19. Nat Commun 13, 4872 (2022).

14. Ferreira, I. A. T. M. et al. Atypical B cells and impaired SARS-CoV-2 neutralisation following booster vaccination in the elderly. 2022.10.13.22281024 Preprint at https://doi.org/10.1101/2022.10.13.22281024 (2022).

15. Madhi, S. A. et al. Durability of ChAdOx1 nCoV-19 (AZD1222) vaccine and hybrid humoral immunity against variants including omicron BA.1 and BA.4 6 months after vaccination (COV005): a post-hoc analysis of a randomised, phase 1b–2a trial. The Lancet Infectious Diseases 23, 295–306 (2023).

16. Abdullahi, A. et al. SARS-COV-2 antibody responses to AZD1222 vaccination in West Africa. Nat Commun 13, 6131 (2022).

17. Shete, A. M. et al. Waning natural and vaccine-induced immunity leading to reinfection with SARS-CoV-2 Omicron variant. Human Vaccines & Immunotherapeutics 18, 2127289 (2022).

18. Sun, P. et al. Antibody Responses to the SARS-CoV-2 Ancestral Strain and Omicron Variants in Moderna mRNA-1273 Vaccinated Active-Duty US Navy Sailors and Marines. The Journal of Infectious Diseases jiad054 (2023) doi:10.1093/infdis/jiad054.

19. Cowling, B. J. et al. Slow Waning of Antibodies Following BNT162b2 as a Third Dose in Adults Who Had Previously Received 2 Doses of Inactivated Vaccine. The Journal of Infectious Diseases 227, 251–255 (2023).

20. Kimura, I. et al. Virological characteristics of the SARS-CoV-2 Omicron BA.2 subvariants, including BA.4 and BA.5. Cell 185, 3992–4007.e16 (2022).

21. Arunachalam, P. S. et al. Durability of immune responses to the booster mRNA vaccination against COVID-19. J Clin Invest (2023) doi:10.1172/JCI167955.

22. Qu, P. et al. Durability of Booster mRNA Vaccine against SARS-CoV-2 BA.2.12.1, BA.4, and BA.5 Subvariants. New England Journal of Medicine 387, 1329–1331 (2022).

23. Cheung, A. K. L. et al. Consistent neutralization of circulating omicron sub-variants by hybrid immunity up to 6 months after booster vaccination. Journal of Medical Virology 95, e28694 (2023).

24. Itamochi, M. et al. Neutralization of Omicron subvariants BA.1 and BA.5 by a booster dose of COVID-19 mRNA vaccine in a Japanese nursing home cohort. Vaccine 41, 2234–2242 (2023).

25. Nair, M. S. et al. Changes in serum-neutralizing antibody potency and breadth post-SARS-CoV-2 mRNA vaccine boost. iScience 26, (2023).

26. Canetti, M. et al. Immunogenicity and efficacy of fourth BNT162b2 and mRNA1273 COVID-19 vaccine doses; three months follow-up. Nat Commun 13, 7711 (2022).

27. Nugent, C. et al. Second monovalent SARS-CoV-2 mRNA booster restores Omicron-specific neutralizing activity in both nursing home residents and health care workers. medRxiv 2023.01.22.23284881 (2023) doi:10.1101/2023.01.22.23284881.

28. Lasrado, N. et al. Waning Immunity Against XBB.1.5 Following Bivalent mRNA Boosters. bioRxiv 2023.01.22.525079 (2023) doi:10.1101/2023.01.22.525079.

29. Jacobsen, H. et al. Diminished neutralization responses towards SARS-CoV-2 Omicron VoC after mRNA or vector-based COVID-19 vaccinations. 2021.12.21.21267898 Preprint at https://doi.org/10.1101/2021.12.21.21267898 (2022).

30. Tada, T. et al. Increased resistance of SARS-CoV-2 Omicron variant to neutralization by vaccine-elicited and therapeutic antibodies. eBioMedicine 78, 103944 (2022).

31. Zheng, H. et al. Disease profile and plasma neutralizing activity of post-vaccination Omicron BA.1 infection in Tianjin, China: a retrospective study. Cell Res 32, 781–784 (2022).

32. Edara, V.-V. et al. mRNA-1273 and BNT162b2 mRNA vaccines have reduced neutralizing activity against the SARS-CoV-2 omicron variant. Cell Reports Medicine 3, 100529 (2022).

33. Garcia-Beltran, W. F. et al. mRNA-based COVID-19 vaccine boosters induce neutralizing immunity against SARS-CoV-2 Omicron variant. Cell 185, 457–466.e4 (2022).

