## Supplementary appendix 1 for "Waning of post-vaccination neutralizing antibody responses against SARS-CoV-2, a systematic literature review and meta-analysis"

| ID | Study First Author | Study population | Naive | | | | | | | | | | Hybrid | | | | | |
|---|---|---|---|---|---|---|---|---|---|---|---|---|---|---|---|---|---|---|
| | | | Primary series | | | | | Boost vaccination | | | | | Primary series | | | | | |
| | | | Vaccine | Number of doses | Time since final dose (months) | WT GMT (95% CI) | BA.1 GMT (95% CI) | Vaccine | Number of doses | Time since final dose (months) | WT GMT (95% CI) | BA.1 GMT (95% CI) | Vaccine | Number of doses | Infecting strain | Time since final dose (months) | WT GMT (95% CI) | BA.1 GMT (95% CI) |
| 24 | Jacobsen | General | BNT+BNT | 2 | 1 | 159.0 (89.3 - 283.0)* | | | | | | | | | | | | |
| | | | BNT+BNT | 2 | 6 | 45.4 (27.2 - 75.8)* | | | | | | | | | | | | |
| 46 | Tada | General | BNT+BNT | 2 | 1 | 878.2 (562.1 - 1371.9)* | | | | | | | BNT+BNT | 2 | n/a | 1 | 3868.0 (2730.7 - 5479.0)* | 215.3 (188.1 - 246.4)* |
| | | | BNT+BNT | 2 | 7.5 | 443.7 (277.0 - 710.8)* | | | | | | | BNT+BNT | 2 | n/a | 7.5 | 2903.8 (1960.0 - 4302.2)* | 125.0 (108.0 - 144.6)* |
| 63 | Lyke | General | | | | | | BNT+BNT+J&J | 3 | 1 | 1090.5 (807.4 - 1473.0) | 358.9 (250.0 - 515.4) | | | | | | |
| | | | | | | | | BNT+BNT+J&J | 3 | 3 | 1444.3 (906.2 - 2302.0) | 123.0 (81.8 - 185.0) | | | | | | |
| | | | | | | | | BNT+BNT+BNT | 3 | 1 | 1305.5 (1044.3 - 1631.9) | 464.1 (341.3 - 631.1) | | | | | | |
| | | | | | | | | BNT+BNT+BNT | 3 | 3 | 886.1 (643.5 - 1220.1) | 131.0 (91.5 - 487.6) | | | | | | |
| | | | | | | | | MOD+MOD+MOD | 3 | 1 | 3383.0 (2673.3 - 4281.2) | 586.8 (426.0 - 808.4) | | | | | | |
| | | | | | | | | MOD+MOD+MOD | 3 | 3 | 1668.0 (1049.0 - 2652.4) | 247.1 (170.3 - 358.4) | | | | | | |
| | | | | | | | | J&J+BNT | 2 | 1 | 1001.6 (784.6 - 1278.7) | 238.8 (173.9 - 327.9) | | | | | | |
| | | | | | | | | J&J+BNT | 2 | 3 | 606.0 (387.5 - 947.6) | 63.6 (46.1 - 87.8) | | | | | | |
| | | | | | | | | J&J+J&J | 2 | 1 | 122.8 (92.0 - 163.8) | 26.9 (19.5 - 37.2) | | | | | | |
| | | | | | | | | J&J+J&J | 2 | 3 | 138.2 (89.3 - 213.9) | 11.2 (8.8 - 14.2) | | | | | | |
| 68 | Walls | General | RNA+RNA | 2 | 0.5 | 3700.0 (n/a) | | | | | | | RNA+RNA | 2 | n/a | 0.5 | 3100.0 (n/a) | 460.0 (n/a) |
| | | | RNA+RNA | 2 | 4 | 1400.0 (n/a) | | | | | | | RNA+RNA | 2 | n/a | 4 | 2600.0 (n/a) | 180.0 (n/a) |
| | | | RNA+RNA | 2 | 6 | 1300.0 (n/a) | | | | | | | RNA+RNA | 2 | n/a | 6.5 | 1800.0 (n/a) | 99.0 (n/a) |
| 70 | Xia | General | BNT+BNT | 2 | 0.5 | 660.5 (466.7 - 934.8)* | | BNT+BNT+BNT | 3 | 1 | 1390.0 (956.0 - 2021.6)* | 353.3 (252.2 - 495.0)* | | | | | | |
| | | | BNT+BNT | 2 | 8 | 68.3 (39.7 - 117.4)* | | BNT+BNT+BNT | 3 | 4 | 819.9 (574.2 - 1170.8)* | 170.9 (113.6 - 257.2)* | | | | | | |
| 127 | Pajon | General | MOD+MOD | 2 | 1 | 1496.0 (916.3 - 2457.3) | | MOD+MOD+MOD | 3 | 1 | 2423.0 (1543.5 - 3750.2) | 850.0 (459.4 - 1610.1) | | | | | | |
| | | | MOD+MOD | 2 | 7 | 193.0 (118.8 - 318.5) | | MOD+MOD+MOD | 3 | 6 | 1067.0 (752.3 - 1543.5) | 136.0 (72.5 - 257.8) | | | | | | |
| | | | MOD+MOD | 2 | 1 | 1165.0 (n/a) | | | | | | | | | | | | |
| | | | MOD+MOD | 2 | 9 | 242.5 (n/a) | | | | | | | | | | | | |
| 149 | Singanallur | General | BNT+BNT | 2 | 0.5 | 100.8 (26.3 - 175.3) | | | | | | | | | | | | |
| | | | BNT+BNT | 2 | 6 | 11.5 (8.3 - 14.6) | | | | | | | | | | | | |
| 152 | Muecksch | General | RNA+RNA | 2 | 1 | 1892.8 (1405.0 - 2550.0)* | | | | | | | | | | | | |
| | | | RNA+RNA | 2 | 5 | 272.2 (190.8 - 388.4)* | | | | | | | | | | | | |
| 217 | Cowling | General | | | | | | INA+INA+BNT | 3 | 1 | 338.0 (n/a) | | | | | | | |
| | | | | | | | | INA+INA+BNT | 3 | 6 | 112.0 (n/a) | | | | | | | |

| ID | Study First Author | Study population | Naive | | | | | | | | | | Hybrid | | | | | |
|---|---|---|---|---|---|---|---|---|---|---|---|---|---|---|---|---|---|---|
| | | | Primary series | | | | | Boost vaccination | | | | | Primary series | | | | | |
| | | | Vaccine | Number of doses | Time since final dose (months) | WT GMT (95% CI) | BA.1 GMT (95% CI) | Vaccine | Number of doses | Time since final dose (months) | WT GMT (95% CI) | BA.1 GMT (95% CI) | Vaccine | Number of doses | Infecting strain | Time since final dose (months) | WT GMT (95% CI) | BA.1 GMT (95% CI) |
| 228 | Behrens | HCW | BNT+BNT | 2 | 1 | 4096.5 (3032.2 - 5534.3)* | | | | | | | | | | | | |
| | | | BNT+BNT | 2 | 7 | 490.8 (337.5 - 713.7)* | | | | | | | | | | | | |
| | | | BNT+BNT | 2 | 9 | 259.6 (190.1 - 354.4)* | | | | | | | | | | | | |
| | | | VAX+VAX | 2 | 0.5 | 335.8 (224.3 - 502.8)* | | | | | | | | | | | | |
| | | | VAX+VAX | 2 | 4 | 169.3 (119.6 - 239.7)* | | | | | | | | | | | | |
| | | | VAX+VAX | 2 | 6 | 49.6 (30.3 - 81.2)* | | | | | | | | | | | | |
| | | | VAX+BNT | 2 | 0.5 | 3104.7 (2425.2 - 3974.6)* | | | | | | | | | | | | |
| | | | VAX+BNT | 2 | 4 | 650.2 (535.7 - 789.2)* | | | | | | | | | | | | |
| | | | VAX+BNT | 2 | 6 | 490.4 (359.7 - 668.6)* | | | | | | | | | | | | |
| 232 | Jin | General | | | | | | COV+COV+Ad5_low | 3 | 1 | 1937.3 (1466.9 - 2558.4) | 52.0 (37.2 - 72.6) | | | | | | |
| | | | | | | | | COV+COV+Ad5_low | 3 | 3 | 530.1 (412.5 - 681.1) | 27.9 (18.8 - 41.3) | | | | | | |
| | | | | | | | | COV+COV+Ad5_low | 3 | 6 | 312.9 (237.7 - 411.8) | | | | | | | |
| | | | | | | | | COV+COV+Ad5_hi | 3 | 1 | 1350.8 (952.6 - 1915.3) | 23.1 (15.7 - 33.9) | | | | | | |
| | | | | | | | | COV+COV+Ad5_hi | 3 | 3 | 457.6 (349.4 - 599.2) | 23.3 (16.2 - 33.3) | | | | | | |
| | | | | | | | | COV+COV+Ad5_hi | 3 | 6 | 251.1 (178.2 - 354.0) | | | | | | | |
| | | | | | | | | COV+COV+COV | 3 | 1 | 73.5 (52.3 - 103.3) | | | | | | | |
| | | | | | | | | COV+COV+COV | 3 | 3 | 20.4 (14.3 - 29.1) | | | | | | | |
| 272 | Zhang | General | | | | | | COV+COV+COV | 3 | 0.5 | 75.4 (61.4 - 92.5) | 8.1 (6.1 - 10.7) | | | | | | |
| | | | | | | | | COV+COV+COV | 3 | 3 | 18.5 (14.9 - 22.9) | 4.7 (4.1 - 5.5) | | | | | | |
| 274 | Kimura | General | | | | | | BNT+BNT+BNT | 3 | 1 | 7210.0 (6340.0 - 8200.0) | | | | | | | |
| | | | | | | | | BNT+BNT+BNT | 3 | 4 | 5530.0 (4330.0 - 6850.0) | | | | | | | |
| 277 | Ferreira | General | VAX+VAX | 2 | 1 | 368.9 (201.2 - 676.5)* | | | | | | | | | | | | |
| | | | VAX+VAX | 2 | 6 | 143.6 (83.8 - 246.1)* | | | | | | | | | | | | |
| 284 | Madhi | General | VAX+VAX | 2 | 1.5 | 451.0 (197.0 - 1035.0) | | | | | | | VAX+VAX | 2 | WT | 1 | 1496.0 (768.0 - 2916.0) | 499.0 (282.0 - 885.0) |
| | | | VAX+VAX | 2 | 6 | 78.0 (19.0 - 329.0) | | | | | | | VAX+VAX | 2 | WT | 1.5 | 1933.0 (1283.0 - 2912.0) | 535.0 (290.0 - 988.0) |
| | | | | | | | | | | | | | VAX+VAX | 2 | WT | 6 | 590.0 (337.0 - 1032.0) | 21.0 (14.0 - 32.0) |
| 292 | Jäger | General | VAX+VAC | 2 | 1 | 325.4 (234.8 - 450.9) | | | | | | | | | | | | |
| | | | VAX+VAC | 2 | 6 | 246.3 (192.7 - 314.6) | | | | | | | | | | | | |
| 130 | Abdullahi | General | VAX+VAX | 2 | 1 | 1495.9 (582.2 - 3843.3)* | | | | | | | VAX+VAX | 2 | Pre-Omicron | 1 | 3127.6 (1969.4 - 4967.0)* | 340.8 (239.8 - 484.2)* |
| | | | VAX+VAX | 2 | 3 | 581.8 (217.3 - 1557.6)* | | | | | | | VAX+VAX | 2 | Pre-Omicron | 3 | 2471.4 (1617.3 - 3776.5)* | 325.7 (235.1 - 451.2)* |

| ID | Study First Author | Study population | Naive | | | | | | | | | | Hybrid | | | | | |
|---|---|---|---|---|---|---|---|---|---|---|---|---|---|---|---|---|---|---|
| | | | Primary series | | | | | Boost vaccination | | | | | Primary series | | | | | |
| | | | Vaccine | Number of doses | Time since final dose (months) | WT GMT (95% CI) | BA.1 GMT (95% CI) | Vaccine | Number of doses | Time since final dose (months) | WT GMT (95% CI) | BA.1 GMT (95% CI) | Vaccine | Number of doses | Infecting strain | Time since final dose (months) | WT GMT (95% CI) | BA.1 GMT (95% CI) |
| 153 | Evans | HCW | RNA+RNA | 2 | 1 | 1558.2 (1159.8 - 2093.4)* | | | | | | | RNA+RNA | 2 | Pre-Omicron | 1 | 6680.4 (2832.7 - 15754.7)* | 94.7 (13.1 - 682.2)* |
| | | | RNA+RNA | 2 | 6 | 91.4 (56.7 - 147.3)* | | | | | | | RNA+RNA | 2 | Pre-Omicron | 6 | 680.7 (152.6 - 3037.1)* | 24.4 (3.1 - 191.8)* |
| 219 | Qu | HCW | | | | | | RNA+RNA+RNA | 3 | 2 | 4448.0 (n/a) | 917.0 (n/a) | | | | | | |
| | | | | | | | | RNA+RNA+RNA | 3 | 5 | 2285.0 (n/a) | 392.0 (n/a) | | | | | | |
| | | | | | | | | RNA+RNA+RNA | 3 | 8 | 1187.0 (n/a) | 257.0 (n/a) | | | | | | |
| 321 | Arunachalam | General | | | | | | RNA+RNA+RNA | 3 | 1 | 1557.3 (1527.2 - 1587.4)* | 127.6 (116.1 - 139.1)* | | | | | | |
| | | | | | | | | RNA+RNA+RNA | 3 | 6 | 386.2 (365.0 - 407.4)* | 29.9 (23.0 - 36.8)* | | | | | | |
| 344 | Wang | General | BBIBP+BBIBP | 2 | 1 | 63.0 (n/a) | | | | | | | | | | | | |
| | | | BBIBP+BBIBP | 2 | 3 | 15.0 (n/a) | | | | | | | | | | | | |
| 365 | Nugent | HCW | | | | | | RNA+RNA+RNA | 3 | 0.5 | 680.0 (467.0 - 990.0) | 114.0 (61.0 - 211.0) | | | | | | |
| | | | | | | | | RNA+RNA+RNA | 3 | 5 | 143.0 (82.0 - 251.0) | 31.0 (17.0 - 56.0) | | | | | | |
| | | Older adults | | | | | | RNA+RNA+RNA | 3 | 0.5 | 611.0 (404.0 - 925.0) | 159.0 (99.0 - 256.0) | | | | | | |
| | | | | | | | | RNA+RNA+RNA | 3 | 5 | 166.0 (77.0 - 174.0) | 27.0 (19.0 - 40.0) | | | | | | |
| 176 | Habermann | General | | | | | | VAC+VAC+BNT | 3 | 1 | 3661.0 (1556.8 - 8404.0) | 729.0 (270.1 - 1889.1) | | | | | | |
| | | | | | | | | VAC+VAC+BNT | 3 | 3 | 1842.0 (919.2 - 3523.0) | 329.0 (129.4 - 814.6) | | | | | | |
| 162 | Canaday | HCW | | | | | | | | | | | BNT+BNT | 2 | Pre-Omicron | 0.5 | 1073.0 (475.0 - 2426.0) | |
| | | | | | | | | | | | | | BNT+BNT | 2 | Pre-Omicron | 8.5 | 39.6 (17.7 - 88.3) | |
| | | Older adults | | | | | | | | | | | BNT+BNT | 2 | Pre-Omicron | 0.5 | 1311.0 (697.0 - 2469.0) | |
| | | | | | | | | | | | | | BNT+BNT | 2 | Pre-Omicron | 8.5 | 34.6 (19.2 - 62.1) | |
| 285 | Shete | n/a | | | | | | | | | | | VAX+VAX | 2 | WT | 2 | 4190.0 (3218.0 - 5542.0) | 212.0 (75.8 - 623.0) |
| | | | | | | | | | | | | | VAX+VAX | 2 | WT | 7 | 1248.0 (713.0 - 2287.0) | 12.0 (1.6 - 97.0) |
| 14 | Gruell | General | BNT+BNT | 2 | 1 | 546.3 (382.9 - 779.5)* | | | | | | | | | | | | |
| | | | BNT+BNT | 2 | 5 | 138.6 (27.2 - 75.8)* | | | | | | | | | | | | |
