## Supplementary appendix 2 for "Waning of post-vaccination neutralizing antibody responses against SARS-CoV-2, a systematic literature review and meta-analysis"

| | | Reference | Tada et al. | | Lyke et al. | | Walls et al. | | Xia et al. | | Lapa et al. | | Pajon et al. | | Abdullahi et al. | | Gilboa et al. | | Singanallur et al. | | Mueksch et al. | |
|---|---|---|---|---|---|---|---|---|---|---|---|---|---|---|---|---|---|---|---|---|---|---|
| **Category** | **Aspect** | **Parameter / explanation** | **Status** | **Impact on reliability** | **Status** | **Impact on reliability** | **Status** | **Impact on reliability** | **Status** | **Impact on reliability** | **Status** | **Impact on reliability** | **Status** | **Impact on reliability** | **Status** | **Impact on reliability** | **Status** | **Impact on reliability** | **Status** | **Impact on reliability** | **Status** | **Impact on reliability** |
| Cohort details | Sample size | **Sample size** *How many samples were included?* | 5-20 | | 21-50 | | 5-20 | | 5-20 | | 5-20 | | 5-20 | | 21-50 | | 21-50 | | 5-20 | | 21-50 | |
| | SARS-CoV-2 infection | **Reported** *Was pre-vaccination COVID-19 considered?* | Yes - and subjects stratified | | Yes - only naive included | | Yes - and subjects stratified | | Not reported and early pandemic setting | | Yes - and subjects stratified | | Yes - only naive included | | Yes - and subjects stratified | | Yes - only naive included | | Not reported and late pandemic setting | | Yes - only naive included | |
| | | **Confirmed** *Previous infection confirmed by NP-ELISA or similar means?* | Yes | | Yes | | Yes | | No / not reported | | Yes | | Yes | | Yes | | Yes | | N. a. | | Yes | |
| | | **Breakthrough cases reported** *Are breakthrough cases reported? Applicable if this is relevant in the context of the study.* | No | | Yes | | Yes | | No | | No | | Yes | | Yes | | Yes | | N. a. | | Yes | |
| | | **Breakthrough cases stratified** *If breakthrough cases occured, did the authors stratify?* | N. a. | | Yes | | Yes | | N. a. | | N. a. | | No | | Yes | | Yes | | N. a. | | N. a. | |
| | Vaccination regimen | **Dosing interval reported** *Do the authors report the dosing interval (if applicable)?* | N. a. | | Yes | | N. a. | | Yes | | N. a. | | Yes | | N. a. | | Yes | | N. a. | | N. a. | |
| | | **Stratified by partial / full immunization** *Do the authors stratiy by partial and full immunization?* | N. a. | | N. a. | | N. a. | | N. a. | | N. a. | | N. a. | | N. a. | | N. a. | | N. a. | | N. a. | |
| | Sample collection period | **≥7 days post last dose** *Were all samples taken at least seven days post final dose? Adjust to "YES" if <10% of samples are taken earlier.* | Yes | | Yes | | Yes | | Yes | | Yes | | Yes | | Yes | | Yes | | Yes | | Yes | |
| | | **Stratified OR 14 d - 4 mo post full immunization** *Are the results stratified OR are all samples taken between 2 weeks and 4 months post final dose? Adjust to "YES" if <20% of sample do not comply.* | Yes | | Yes | | Yes | | Yes | | Yes | | Yes | | Yes | | Yes | | Yes | | Yes | |
| | Demographic characterization | **Age distribution reported** *Is the age distribution (range) of all subjects reported?* | Yes | | Yes | | Yes | | Yes | | Yes | | Yes | | No | | Yes | | Yes | | Yes | |
| | | **Stratified by age group (<18 or 18 - 59 or ≥60)** *Adjust to "YES" if <20% of samples belong to different age groups* | Yes | | No | | No | | No | | Yes | | No | | No | | No | | Yes | | Yes | |
| | | **Sex distribution reported** *Is the sex distribution of all subjects reported?* | Yes | | Yes | | Yes | | Yes | | Yes | | Yes | | No | | Yes | | Yes | | Yes | |
| | | **Stratified by sex or equal sex distribution** *Equal sex distribution: 50% ±10% per sex. Adjust to "YES" if <20% of samples were not stratified.* | Equal sex distribution | | Equal sex distribution | | Not stratified nor equally distributed | | Not stratified nor equally distributed | | Equal sex distribution | | Equal sex distribution | | Not stratified nor equally distributed | | Not stratified nor equally distributed | | Not stratified nor equally distributed | | Equal sex distribution | |
| | | **Cohort selection unbiased** *If only a subgroup of the initial study cohort was analyzed, did the cohort selection happen unbiased (no pre-selection of high-titre reponders etc)?* | Yes | | Yes | | Yes | | Yes | | Yes | | Yes | | Yes | | Yes | | Yes | | Yes | |
| | | **Study period and geographic location reported** *Applicable if SARS-CoV-2 infections occured and variant identification & distribution is not reported for the cohort.* | Yes | | N. a. | | Yes | | Yes | | N. a. | | N. a. | | Yes | | N. a. | | N. a. | | N. a. | |
| | | **Variant prevalence reported** *Applicable if any SARS-CoV-2 infections occured.* | Yes | | N. a. | | Yes | | N. a. | | N. a. | | N. a. | | Yes | | N. a. | | N. a. | | N. a. | |
| | | **Stratified by variant prevalence** *Applicable if SARS-CoV-2 infections with multiple variants occured.* | No | | N. a. | | Yes | | N. a. | | N. a. | | N. a. | | No | | N. a. | | N. a. | | N. a. | |
| | Clinical characterization | **Reported** *Is any relevant clinical characterization reported? Applicable if at least one third of the study cohort is likely or known to have clinical conditions that might affect immunity.* | Yes | | N. a. | | No | | N. a. | | N. a. | | N. a. | | N. a. | | N. a. | | N. a. | | N. a. | |
| | | **Stratifed by immunocompromised** *Applicable if clinical characterization applies. Adjust to "YES" if <20% of samples were not stratified.* | Yes | | N. a. | | N. a. | | N. a. | | N. a. | | N. a. | | N. a. | | N. a. | | N. a. | | N. a. | |
| Assay details | Protocol | **Assay type reported** *Is the precise assay type and endpoint reported (pseudovirus vs life virus, NT20, NT50, NT80 etc)?* | Yes | | Yes | | Yes | | Yes | | Yes | | Yes | | Yes | | Yes | | | | Yes | |
| | | **Precise protocol reported** *Do the authors provide a precise protocol for the neutralization assay within the manuscript?* | No | | Yes | | Yes | | Yes | | Yes | | Yes | | Yes | | Yes | | Yes | | Yes | |
| | Live virus strain (if applicable) | **Virus lineage reported** *Applicable if live virus neutralization was performed.* | N. a. | | N. a. | | N. a. | | Yes | | Yes | | Yes | | N. a. | | No | | Yes | | N. a. | |
| | | **Sequence confirmation by sequencing** *Applicable if live virus neutralization was performed.* | N. a. | | N. a. | | N. a. | | Yes | | No | | Yes | | N. a. | | No | | Yes | | N. a. | |
| | Pseudo virus strain (if applicable) | **Construct details reported** *Applicable if pseudovirus neutralization was performed.* | No | | Yes | | No | | N. a. | | N. a. | | Yes | | No | | N. a. | | N. a. | | Yes | |
| | | **All variant-associated spike mutations** *Applicable if pseudovirus neutralization was performed. Are all variant-associated spike mutations included to the pseudovirus?* | N. a. | | Yes | | No | | N. a. | | N. a. | | Yes | | N. a. | | N. a. | | N. a. | | Yes | |
| | | **Sequence confirmation by sequencing** *Applicable if pseudovirus neutralization was performed.* | N. a. | | Yes | | No | | N. a. | | N. a. | | No | | N. a. | | N. a. | | N. a. | | No | |
| | Assay standardization | **Virus titre reported and consistent** *Are virus titres used for neutralization assays reported and if so: consistent and with small input variance?* | Consistent and with small variance | | Not reported | | Not reported | | Consistent and with small variance | | Consistent and with small variance | | Consistent and with small variance | | Not reported | | Consistent and with small variance | | Consistent and with small variance | | Not reported | |
| | | **Error in titre reported by back titration** *Was the virus titre used for neutralization assays confirmed by the authors by back-titration or similar means?* | No | | No | | No | | No | | No | | No | | No | | No | | Yes | | No | |
| | | **WHO IS antibody used** *WHO international standard antibody used for standardization?* | No | | | | | | | | | | | | | | | | | | | |
| | | **Details on cell culture reported** *Are precise details on cell culture reported (cell culture conditions, maximum passage number etc)?* | Yes | | Yes | | Yes | | Yes | | Yes | | No | | No | | No | | Yes | | | |
| Data | Data reporting | **Raw data reported** *Raw data for neutralization titres reported?* | Yes | | No | | No | | Yes | | No | | No | | No | | No | | No | | Yes | |
| | | **Reference virus is appropriate (non VoI/VoC)** *Is the reference virus used for calculating variant-specific fold-changes reasonable (non VoI/VoC)?* | Yes | | Yes | | Yes | | Yes | | Yes | | Yes | | Yes | | Yes | | Yes | | Yes | |
| | | **Data shown as individual values with statistics** *Are individual data points and apropriate statistics provided?* | Yes | | Yes | | Yes | | Yes | | Yes | | Yes | | Yes | | Yes | | Yes | | Yes | |
| | | | **Overall risk of low reliability:** | UNCLEAR | **Overall risk of low reliability:** | UNCLEAR | **Overall risk of low reliability:** | UNCLEAR | **Overall risk of low reliability:** | MEDIUM | **Overall risk of low reliability:** | MEDIUM | **Overall risk of low reliability:** | HIGH | **Overall risk of low reliability:** | UNCLEAR | **Overall risk of low reliability:** | UNCLEAR | **Overall risk of low reliability:** | UNCLEAR | **Overall risk of low reliability:** | UNCLEAR |

| | | Reference | Evans et al. | | Canaday et al. | | Cowling et al. | | Behrens et al. | | Jin et al. | | Zhang et al. | | Kimura et al. | | Ferreira et al. | | Madhi et al. | | Jaeger et al. | |
|---|---|---|---|---|---|---|---|---|---|---|---|---|---|---|---|---|---|---|---|---|---|---|
| **Category** | **Aspect** | **Parameter / explanation** | **Status** | **Impact on reliability** | **Status** | **Impact on reliability** | **Status** | **Impact on reliability** | **Status** | **Impact on reliability** | **Status** | **Impact on reliability** | **Status** | **Impact on reliability** | **Status** | **Impact on reliability** | **Status** | **Impact on reliability** | **Status** | **Impact on reliability** | **Status** | **Impact on reliability** |
| Cohort details | Sample size | **Sample size**<br>*How many samples were included?* | <5 | | 5-20 | | 21-50 | | 21-50 | | 21-50 | | 21-50 | | 5-20 | | 21-50 | | 5-20 | | 5-20 | |
| | SARS-CoV-2 infection | **Reported**<br>*Was pre-vaccination COVID-19 considered?* | Yes - and subjects stratified | | Yes - and subjects stratified | | Yes - only naive included | | Yes - only naive included | | Yes - only naive included | | Yes - only naive included | | Yes - only naive included | | Yes - only naive included | | Yes - and subjects stratified | | Yes - only naive included | |
| | | **Confirmed**<br>*Previous infection confirmed by NP-ELISA or similar means?* | Yes | | Yes | | No / not reported | | Yes | | No / not reported | | No / not reported | | No / not reported | | Yes | | Yes | | Yes | |
| | | **Breakthrough cases reported**<br>*Are breakthrough cases reported? Applicable if this is relevant in the context of the study.* | Yes | | Yes | | No | | Yes | | No | | No | | No | | Yes | | Yes | | Yes | |
| | | **Breakthrough cases stratified**<br>*If breakthrough cases occured, did the authors stratify?* | Yes | | Yes | | No | | Yes | | N. a. | | N. a. | | N. a. | | N. a. | | Yes | | N. a. | |
| | Vaccination regimen | **Dosing interval reported**<br>*Do the authors report the dosing interval (if applicable)?* | N. a. | | N. a. | | Yes | | Yes | | Yes | | Yes | | N. a. | | N. a. | | N. a. | | N. a. | |
| | | **Stratified by partial / full immunization**<br>*Do the authors stratiy by partial and full immunization?* | N. a. | | Yes | | Yes | | Yes | | N. a. | | Yes | | Yes | | Yes | | Yes | | Yes | |
| | Sample collection period | **≥7 days post last dose**<br>*Were all samples taken at least seven days post final dose?*<br>*Adjust to "YES" if <10% of samples are taken earlier.* | Yes | | Yes | | Yes | | Yes | | Yes | | Yes | | Yes | | Yes | | Yes | | Yes | |
| | | **Stratified OR 14 d - 4 mo post full immunization**<br>*Are the results stratified OR are all samples taken between 2 weeks and 4 months post final dose?*<br>*Adjust to "YES" if <20% of sample do not comply.* | Yes | | Yes | | Yes | | Yes | | Yes | | Yes | | Yes | | Yes | | Yes | | Yes | |
| | Demographic characterization | **Age distribution reported**<br>*Is the age distribution (range) of all subjects reported?* | Yes | | Yes | | Yes | | Yes | | Yes | | Yes | | Yes | | Yes | | Yes | | Yes | |
| | | **Stratified by age group (<18 or 18 - 59 or ≥60)**<br>*Adjust to "YES" if <20% of samples belong to different age groups* | Yes | | Yes | | No | | Yes | | Yes | | Yes | | Yes | | Yes | | Yes | | Yes | |
| | | **Sex distribution reported**<br>*Is the sex distribution of all subjects reported?* | Yes | | Yes | | Yes | | Yes | | Yes | | Yes | | Yes | | Yes | | Yes | | Yes | |
| | | **Stratified by sex or equal sex distribution**<br>*Equal sex distribution: 50% ±10% per sex.*<br>*Adjust to "YES" if <20% of samples were not stratified.* | Equal sex distribution | | Not stratified nor equally distributed | | Not stratified nor equally distributed | | Not stratified nor equally distributed | | Not stratified nor equally distributed | | Equal sex distribution | | Equal sex distribution | | Not stratified nor equally distributed | | Equal sex distribution | | Not stratified nor equally distributed | |
| | | **Cohort selection unbiased**<br>*If only a subgroup of the initial study cohort was analyzed, did the cohort selection happen unbiased (no pre-selection of high-titre reponders etc)?* | Yes | | Yes | | Yes | | Yes | | Yes | | Yes | | Yes | | Yes | | Yes | | Yes | |
| | | **Study period and geographic location reported**<br>*Applicable if SARS-CoV-2 infections occured and variant identification & distribution is not reported for the cohort.* | Yes | | Yes | | N. a. | | N. a. | | N. a. | | N. a. | | N. a. | | N. a. | | Yes | | N. a. | |
| | | **Variant prevalence reported**<br>*Applicable if any SARS-CoV-2 infections occured.* | No | | No | | N. a. | | N. a. | | N. a. | | N. a. | | N. a. | | N. a. | | No | | N. a. | |
| | | **Stratified by variant prevalence**<br>*Applicable if SARS-CoV-2 infections with multiple variants occured.* | No | | No | | N. a. | | N. a. | | N. a. | | N. a. | | N. a. | | N. a. | | No | | | |
| | Clinical characterization | **Reported**<br>*Is any relevant clinical characterization reported?*<br>*Applicable if at least one third of the study cohort is likely or known to have clinical conditions that might affect immunity.* | N. a. | | No | | Yes | | N. a. | | N. a. | | N. a. | | N. a. | | No | | N. a. | | N. a. | |
| | | **Stratifed by immunocompromised**<br>*Applicable if clinical characterization applies.*<br>*Adjust to "YES" if <20% of samples were not stratified.* | N. a. | | | | No | | N. a. | | N. a. | | N. a. | | N. a. | | N. a. | | N. a. | | N. a. | |
| Assay details | Protocol | **Assay type reported**<br>*Is the precise assay type and endpoint reported (pseudovirus vs life virus, NT20, NT50, NT80 etc)?* | Yes | | Yes | | Yes | | Yes | | Yes | | Yes | | Yes | | Yes | | Yes | | Yes | |
| | | **Precise protocol reported**<br>*Do the authors provide a precise protocol for the neutralization assay within the manuscript?* | Yes | | No | | Yes | | No | | Yes | | No | | Yes | | Yes | | Yes | | Yes | |
| | Live virus strain (if applicable) | **Virus lineage reported**<br>*Applicable if live virus neutralization was performed.* | N. a. | | N. a. | | No | | N. a. | | Yes | | Yes | | N. a. | | N. a. | | N. a. | | No | |
| | | **Sequence confirmation by sequencing**<br>*Applicable if live virus neutralization was performed.* | N. a. | | N. a. | | No | | N. a. | | No | | No | | N. a. | | N. a. | | N. a. | | N. a. | |
| | Pseudo virus strain (if applicable) | **Construct details reported**<br>*Applicable if pseudovirus neutralization was performed.* | No | | Yes | | N. a. | | No | | N. a. | | N. a. | | Yes | | Yes | | Yes | | N. a. | |
| | | **All variant-associated spike mutations**<br>*Applicable if pseudovirus neutralization was performed.*<br>*Are all variant-associated spike mutations included to the pseudovirus?* | N. a. | | Yes | | N. a. | | N. a. | | N. a. | | N. a. | | N. a. | | N. a. | | Yes | | N. a. | |
| | | **Sequence confirmation by sequencing**<br>*Applicable if pseudovirus neutralization was performed.* | N. a. | | No | | N. a. | | No | | N. a. | | N. a. | | Yes | | No | | No | | N. a. | |
| | Assay standardization | **Virus titre reported and consistent**<br>*Are virus titres used for neutralization assays reported and if so: consistent and with small input variance?* | Not reported | | Not consistent or with high variance | | Consistent and with small variance | | Not reported | | Consistent and with small variance | | Not reported | | Consistent and with small variance | | Not reported | | Not reported | | Consistent and with small variance | |
| | | **Error in titre reported by back titration**<br>*Was the virus titre used for neutralization assays confirmed by the authors by back-titration or similar means?* | No | | No | | No | | No | | No | | No | | No | | No | | No | | No | |
| | | **WHO IS antibody used**<br>*WHO international standard antibody used for standardization?* | | | | | | | | | | | | | | | | | | | | |
| | | **Details on cell culture reported**<br>*Are precise details on cell culture reported*<br>*(cell culture conditions, maximum passage number etc)?* | Yes | | No | | No | | No | | No | | No | | Yes | | No | | No | | Yes | |
| Data | Data reporting | **Raw data reported**<br>*Raw data for neutralization titres reported?* | Yes | | No | | No | | Yes | | No | | No | | No | | No | | No | | Yes | |
| | | **Reference virus is appropriate (non VoI/VoC)**<br>*Is the reference virus used for calculating variant-specific fold-changes reasonable (non VoI/VoC)?* | Yes | | Yes | | Yes | | Yes | | Yes | | Yes | | Yes | | Yes | | Yes | | Yes | |
| | | **Data shown as individual values with statistics**<br>*Are individual data points and apropriate statistics provided?* | Yes | | Yes | | No | | Yes | | Yes | | Yes | | Yes | | Yes | | Yes | | Yes | |
| | | | **Overall risk of low reliability:** | HIGH | **Overall risk of low reliability:** | HIGH | **Overall risk of low reliability:** | HIGH | **Overall risk of low reliability:** | UNCLEAR | **Overall risk of low reliability:** | MEDIUM | **Overall risk of low reliability:** | UNCLEAR | **Overall risk of low reliability:** | MEDIUM | **Overall risk of low reliability:** | UNCLEAR | **Overall risk of low reliability:** | UNCLEAR | **Overall risk of low reliability:** | UNCLEAR |

| | | Reference | Qu et al. | | Arunachalam et al. | | Wang et al. | | Nugent et al. | | Habermann et al. | | Shete et al. | | Gruell et al. | | Jacobsen et al. | |
|---|---|---|---|---|---|---|---|---|---|---|---|---|---|---|---|---|---|---|
| **Category** | **Aspect** | **Parameter / explanation** | **Status** | **Impact on reliability** | **Status** | **Impact on reliability** | **Status** | **Impact on reliability** | **Status** | **Impact on reliability** | **Status** | **Impact on reliability** | **Status** | **Impact on reliability** | **Status** | **Impact on reliability** | **Status** | **Impact on reliability** |
| Cohort details | Sample size | **Sample size**<br>*How many samples were included?* | 21-50 | | 21-50 | | 21-50 | | 5-20 | | 21-50 | | 5-20 | | 21-50 | | 21-50 | |
| | SARS-CoV-2 infection | **Reported**<br>*Was pre-vaccination COVID-19 considered?* | Yes - and subjects stratified | | Yes - and subjects stratified | | Yes - and subjects stratified | | Yes - only naive included | | Yes - only naive included | | Yes - and subjects stratified | | Yes - only naive included | | Yes - only naive included | |
| | | **Confirmed**<br>*Previous infection confirmed by NP-ELISA or similar means?* | No / not reported | | Yes | | No / not reported | | No / not reported | | No / not reported | | Yes | | Yes | | Yes | |
| | | **Breakthrough cases reported**<br>*Are breakthrough cases reported? Applicable if this is relevant in the context of the study.* | Yes | | Yes | | Yes | | Yes | | Yes | | Yes | | N. a. | | Yes | |
| | | **Breakthrough cases stratified**<br>*If breakthrough cases occured, did the authors stratify?* | No | | N. a. | | N. a. | | N. a. | | Yes | | Yes | | N. a. | | N. a. | |
| | Vaccination regimen | **Dosing interval reported**<br>*Do the authors report the dosing interval (if applicable)?* | N. a. | | Yes | | Yes | | N. a. | | Yes | | N. a. | | N. a. | | Yes | |
| | | **Stratified by partial / full immunization**<br>*Do the authors stratiy by partial and full immunization?* | Yes | | Yes | | Yes | | Yes | | Yes | | N. a. | | N. a. | | Yes | |
| | Sample collection period | **≥7 days post last dose**<br>*Were all samples taken at least seven days post final dose?*<br>*Adjust to "YES" if <10% of samples are taken earlier.* | Yes | | Yes | | Yes | | Yes | | Yes | | Yes | | Yes | | Yes | |
| | | **Stratified OR 14 d - 4 mo post full immunization**<br>*Are the results stratified OR are all samples taken between 2 weeks and 4 months post final dose?*<br>*Adjust to "YES" if <20% of sample do not comply.* | Yes | | Yes | | Yes | | Yes | | Yes | | Yes | | Yes | | Yes | |
| | Demographic characterization | **Age distribution reported**<br>*Is the age distribution (range) of all subjects reported?* | Yes | | Yes | | Yes | | Yes | | Yes | | No | | Yes | | Yes | |
| | | **Stratified by age group (<18 or 18 - 59 or ≥60)**<br>*Adjust to "YES" if <20% of samples belong to different age groups* | Yes | | No | | Yes | | No | | No | | Not reported | | No | | Yes | |
| | | **Sex distribution reported**<br>*Is the sex distribution of all subjects reported?* | Yes | | Yes | | Yes | | Yes | | Yes | | No | | Yes | | Yes | |
| | | **Stratified by sex or equal sex distribution**<br>*Equal sex distribution: 50% ±10% per sex.*<br>*Adjust to "YES" if <20% of samples were not stratified.* | Not stratified nor equally distributed | | Not stratified nor equally distributed | | Not stratified nor equally distributed | | Stratified | | Not stratified nor equally distributed | | Not stratified nor equally distributed | | Equal sex distribution | | Not stratified nor equally distributed | |
| | | **Cohort selection unbiased**<br>*If only a subgroup of the initial study cohort was analyzed, did the cohort selection happen unbiased (no pre-selection of high-titre reponders etc)?* | Yes | | Yes | | Yes | | Yes | | Yes | | Yes | | Yes | | Yes | |
| | | **Study period and geographic location reported**<br>*Applicable if SARS-CoV-2 infections occured and variant identification & distribution is not reported for the cohort.* | Yes | | N. a. | | N. a. | | N. a. | | N. a. | | Yes | | N. a. | | N. a. | |
| | | **Variant prevalence reported**<br>*Applicable if any SARS-CoV-2 infections occured.* | Yes | | N. a. | | N. a. | | N. a. | | N. a. | | Yes | | N. a. | | N. a. | |
| | | **Stratified by variant prevalence**<br>*Applicable if SARS-CoV-2 infections with multiple variants occured.* | No | | N. a. | | N. a. | | N. a. | | N. a. | | No | | N. a. | | N. a. | |
| | Clinical characterization | **Reported**<br>*Is any relevant clinical characterization reported?*<br>*Applicable if at least one third of the study cohort is likely or known to have clinical conditions that might affect immunity.* | N. a. | | N. a. | | N. a. | | No | | Yes | | No | | Yes | | Yes | |
| | | **Stratifed by immunocompromised**<br>*Applicable if clinical characterization applies.*<br>*Adjust to "YES" if <20% of samples were not stratified.* | N. a. | | N. a. | | N. a. | | N. a. | | Yes | | N. a. | | Yes | | Yes | |
| Assay details | Protocol | **Assay type reported**<br>*Is the precise assay type and endpoint reported (pseudovirus vs life virus, NT20, NT50, NT80 etc)?* | Yes | | Yes | | Yes | | Yes | | Yes | | No | | Yes | | Yes | |
| | | **Precise protocol reported**<br>*Do the authors provide a precise protocol for the neutralization assay within the manuscript?* | No | | Yes | | Yes | | Yes | | Yes | | No | | Yes | | Yes | |
| | Live virus strain (if applicable) | **Virus lineage reported**<br>*Applicable if live virus neutralization was performed.* | N. a. | | Yes | | N. a. | | N. a. | | N. a. | | No | | N. a. | | N. a. | |
| | | **Sequence confirmation by sequencing**<br>*Applicable if live virus neutralization was performed.* | N. a. | | Yes | | N. a. | | N. a. | | N. a. | | N. a. | | N. a. | | N. a. | |
| | Pseudo virus strain (if applicable) | **Construct details reported**<br>*Applicable if pseudovirus neutralization was performed.* | No | | N. a. | | Yes | | No | | Yes | | N. a. | | Yes | | Yes | |
| | | **All variant-associated spike mutations**<br>*Applicable if pseudovirus neutralization was performed.*<br>*Are all variant- associated spike mutations included to the pseudovirus?* | N. a. | | N. a. | | Yes | | N. a. | | Yes | | N. a. | | Yes | | Yes | |
| | | **Sequence confirmation by sequencing**<br>*Applicable if pseudovirus neutralization was performed.* | No | | N. a. | | Yes | | N. a. | | Yes | | N. a. | | Yes | | Yes | |
| | Assay standardization | **Virus titre reported and consistent**<br>*Are virus titres used for neutralization assays reported and if so: consistent and with small input variance?* | Not reported | | Consistent and with small variance | | Consistent and with small variance | | Not consistent or with high variance | | Consistent and with small variance | | Not reported | | Not reported | | Consistent and with small variance | |
| | | **Error in titre reported by back titration**<br>*Was the virus titre used for neutralization assays confirmed by the authors by back-titration or similar means?* | No | | No | | No | | No | | No | | No | | No | | Yes | |
| | | **WHO IS antibody used**<br>*WHO international standard antibody used for standardization?* | | | | | | | | | | | | | | | | |
| | | **Details on cell culture reported**<br>*Are precise details on cell culture reported*<br>*(cell culture conditions, maximum passage number etc)?* | No | | Yes | | Yes | | No | | No | | No | | Yes | | Yes | |
| Data | Data reporting | **Raw data reported**<br>*Raw data for neutralization titres reported?* | No | | No | | No | | No | | No | | No | | Yes | | Yes | |
| | | **Reference virus is appropriate (non VoI/VoC)**<br>*Is the reference virus used for calculating variant-specific fold-changes reasonable (non VoI/VoC)?* | Yes | | Yes | | Yes | | Yes | | Yes | | Yes | | Yes | | Yes | |
| | | **Data shown as individual values with statistics**<br>*Are individual data points and apropriate statistics provided?* | Yes | | Yes | | No | | Yes | | Yes | | Yes | | Yes | | Yes | |
| | | | **Overall risk of low reliability:** | HIGH | **Overall risk of low reliability:** | LOW | **Overall risk of low reliability:** | LOW | **Overall risk of low reliability:** | HIGH | **Overall risk of low reliability:** | LOW | **Overall risk of low reliability:** | UNCLEAR | **Overall risk of low reliability:** | UNCLEAR | **Overall risk of low reliability:** | LOW |
